# Embedding Telerehabilitation in Practice: Adapting NeuroRehabilitation OnLine (NROL) for a New Context Through Co-Production and Implementation Science Knowledge

**DOI:** 10.1101/2025.10.13.25337875

**Authors:** Suzanne Ackerley, Amy Bastow, Ruth Witham, Louise Jackson, Adam Partington, Helen Vernon, Louise A Connell

**Author notes:** CORRESPONDING AUTHOR: Prof. Louise A Connell, PhD, Health Innovation Campus, Lancaster University, LA14AT.

## Abstract

**Background:** Delivering recommended intensity of neurorehabilitation remains a challenge, with telerehabilitation offering one solution. NeuroRehabilitation OnLine (NROL) is a multidisciplinary, group-based telerehabilitation model embedded within one region in England, showing promise and alignment with healthcare strategic priorities. The importance of scaling successful evidence-informed practices is recognised, however careful adaptation is required to ensure contextual fit and sustainability. This study describes the adaptation of NROL for implementation in a new context.

**Methods:** The four-step ADAPT guidance was applied with previously identified implementation strategies to guide the adaptation of NROL from Region 1 to Region 2. Adaptation activities were co-produced. Contextual factors were detailed using the Consolidated Framework for Implementation Research and Intervention Sustainability Assessment Tool. The adapted innovation was described using the TIDieR-Rehab checklist.

**Results:** NROL was successfully adapted for Region 2. Step 1 confirmed strategic fit but identified barriers including workforce, infrastructure and resource. Led by an adaptation team and supported by a learning collaborative, Step 2 responded to barriers, retaining core components while tailoring role configuration and materials. Step 3 demonstrated feasibility and acceptability through piloting and phased integration, and improved fit within service pathways. Step 4 focused on sustainment, supported by training, stakeholder engagement, and reporting.

**Discussion:** This study offers a transferable approach for scale-up, providing an example of context-sensitive adaptation to a new region, demonstrating how frameworks and co-production can support the adaptation of telerehabilitation models. This example has potential wider use for researchers and implementers tasked with delivering impact, though highlights the effort and resource needed.

**PLAIN LANGUAGE SUMMARY:** Stroke and neurological rehabilitation services often struggle to provide enough therapy due to staffing and system challenges. Digital delivery of rehabilitation offers one solution.

NeuroRehabilitation OnLine (NROL) is a programme that offers live, group therapy and peer-support remotely for people recovering neurological conditions. It has been delivered within one region of England, showing promise and alignment with healthcare priorities. The importance of spreading successful, evidenced practices is recognised, but requires careful consideration to ensure they fit well and can be continued in the new location. This study explains how NROL was adapted for use in a second region.

The four-step process called the ADAPT guidance was used with previously identified practical strategies to guide changes to NROL for Region 2. Relevant staff from both regions worked together to ensure the programme met local needs. Evidenced tools were used to understand what made the new region different and to describe the adapted programme clearly.

NROL was successfully adapted for Region 2. Step 1 confirmed NROL alignment with healthcare priorities but highlighted challenges including limited staffing and resource. Led by a small team, Step 2 responded to challenges. The core parts of NROL were kept the same, like live group therapy and technology support, but changes were made to the staffing structures and materials. Step 3 tested the changes with small pilot groups and used feedback to improve the programme before wider roll-out and refinement. Step 4 focused on maintaining and reporting the adapted programme.

This work offers a transferable approach for spreading evidenced health innovations, providing an example of local needs driving relevant change, and demonstrating how tools and teamwork can support the adaptation process of digital health programmes. This example has potential wider use for researchers and clinical staff tasked with delivering change, though highlights the effort and resource needed.

## INTRODUCTION

Delivering recommended intensity of neurorehabilitation remains a persistent global challenge,^1–3^ with workforce shortages, geographic disparities, and fragmented care pathways. Telerehabilitation may help address these gaps, offering comparable outcomes to face-to-face therapy and can increase access to specialist rehabilitation through flexible, remote delivery.^4, 5^

NeuroRehabilitation OnLine (NROL) provides real-time, multidisciplinary, group-based, telerehabilitation with dedicated technology assistance.^6^ It combines targeted therapy with peer-support aiming to enhance access and opportunity for neurorehabilitation while making efficient use of the workforce. Recent publications provide a comprehensive description of NROL’s components,^6^ and successful integration within the National Health Service (NHS) stroke and neurorehabilitation services in one region of England (Lancashire and South Cumbria).^7, 8^ These highlight NROL’s alignment with national guidelines,^9, 10^ healthcare strategic priorities,^11–13^ and its adaptability to local contexts.

Scaling evidence-informed practices, like NROL, is increasingly recognised as a strategic objective for health systems. The World Health Organization reports scale-up as “the deliberate effort to increase the impact of successfully tested health interventions so as to benefit more people and to foster policy and program development on a lasting basis”.^14^ Adaptation is a key part of successful scale-up, with careful attention required to how well an intervention will fit and function in the new context, including considering infrastructure, workforce, readiness, patient needs and sustainment. Despite the importance of reporting intervention adaptations, a recent review found many studies lack description in full.^15^ The ADAPT guidance was developed to provide a systematic, stepwise approach to support structured and transparent adaptation.^16^ Alongside ADAPT, TIDieR checklists provide structured tools for describing interventions in sufficient detail to support replication and fidelity.^17–19^ Despite their availability, to the authors knowledge no studies have formally applied both ADAPT and TIDieR comprehensively to describe innovation adaptation in the context of telehealth.

Combining complementary implementation science knowledge with the ADAPT guidance can help navigate the complexities of transferring interventions across diverse health systems.^16, 20^ The value of concurrent use of implementation science knowledge was also highlighted by Connell et al. (2025), using NROL as an example.^21^ They used the ADAPT guidance and the Consolidated Framework for Implementation Research (CFIR),^22^ in conjunction with the updated Medical Research Council framework for the development and evaluation of complex interventions^23^ to develop, adapt, implement and evaluate NROL as a scalable regional innovation. Of note, incorporation of the CFIR enabled identification of Expert Recommendations for Implementing Change (ERIC) implementation strategies (through the CFIR-ERIC matching tool) to support implementation efforts, including for the adaptation phase.

Building on prior work, this paper focuses on adapting NROL for delivery in a new context, a second NHS region within England (Cheshire and Merseyside). Using the ADAPT guidance and TIDieR-Rehab checklist, the adaptation process is documented along with contextual considerations and applied implementation strategies. This publication aims to offer considerations for therapy staff, service leads, and digital health implementers seeking to adapt or scale other telehealth models.

## METHODS

The process was iterative, and grounded in co-production principles with the adaptation of NROL applying complementary implementation science knowledge. Ethical approval was not required for this descriptive study.

### Stakeholder co-production

A key feature of NROL’s development, implementation and adaptation has been co-production, with key stakeholders (including therapy staff, service leads, patients, clinical-academics, and members of the stroke and neurorehabilitation community) actively shaping the innovation. This collaborative model supports contextual relevance, clinician buy-in, and patient engagement; factors known to influence implementation success and sustainability.^24^

A coalition was built between Region 2 (Cheshire and Merseyside) and Region 1 (Lancashire and South Cumbria), facilitated by established connectivity between their regional stroke networks and a shared purpose to improve neurorehabilitation provision, with NROL identified as one possible solution. The adaptation of NROL for Region 2 was shaped by co-production principles emphasising shared power and mutual respect.^25, 26^ Individuals contributed their perspectives and skills with all forms of knowledge valued equally. Fostering reciprocal relationships, prioritising relationship-building and maintaining open communication, ensured the project met the needs of all stakeholders.

#### Positionality statement

The author team comprises clinical-academic staff, and operational, service and regional leads, embedded within the stroke and neurorehabilitation services across both regions involved in this study. Our direct involvement in the development, delivery, and adaptation of NROL provided deep contextual insight and facilitated co-production. We acknowledge that our dual roles, as implementers and evaluators, may have influenced interpretation. To mitigate bias, diverse stakeholders were engaged throughout and structured frameworks used to guide and document the process.

### Process for adapting NROL

Adaptation of NROL in Region 2 used a multi-tool approach, with selection of tools informed by complementary strengths in structuring adaptation processes; capturing contextual influences, ensuring transparent reporting, and fit within the larger programme of work previously described.^21^

The ADAPT guidance was the overarching tool.^16^ NROL was adapted to new contexts informed by four key steps, as outlined in Figure 1. This guidance was particularly suited to the current project, which aimed to scale-up an existing innovation to a new region while maintaining fidelity to its core components. Adaptation was supported using previously identified ERIC implementation strategies.^21^ ERIC strategies help inform actions to support effective implementation.^27^ Strategies focused on stakeholder interrelationships, training and education, and evaluation and iteration.^28^

**Figure 1.**
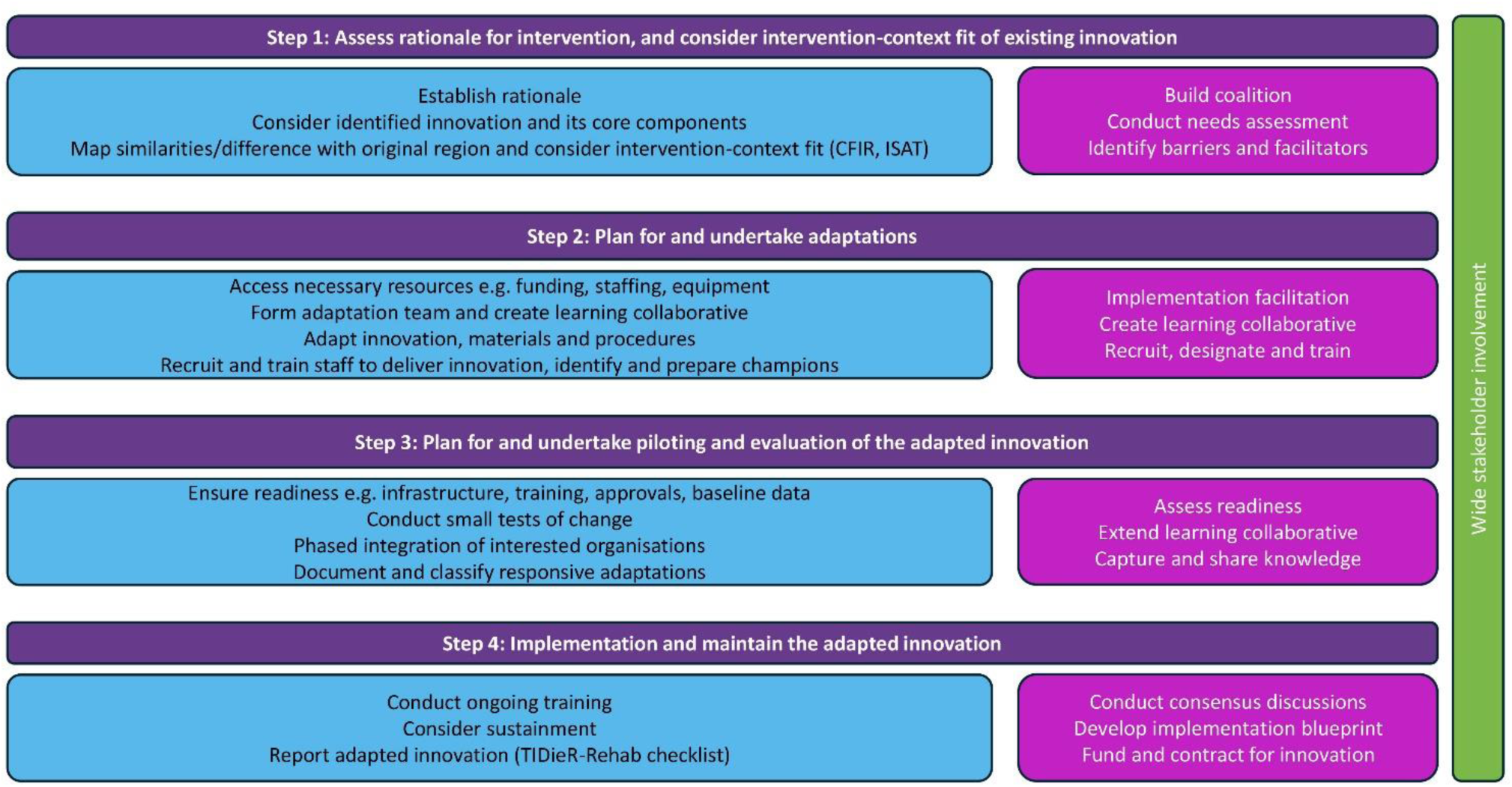
Process for adapting NeuroRehabilitation OnLine (NROL) to new contexts, informed by the ADAPT guidance,^16^ Consolidated Framework for Implementation research (CFIR),^22^ Intervention Scalability Assessment Tool (ISAT),^29^ and reported with the TIDieR-Rehab checklist.^18^ Purple boxes = Steps of the ADAPT guidance. Blue boxes = Expected outcomes from each step. Pink boxes = ERIC implementation strategies applied.^21^

In Step 1, the CFIR and ISAT supported contextual analysis. CFIR offered a comprehensive lens to systematically detail and compare contextual factors (innovation characteristics, outer setting, inner setting, characteristics of individuals) across regions.^22^ The ISAT provided a structured assessment of context-specific factors influencing readiness for scale-up, covering 2 parts: A-Background contextual information and B-Implementation and sustainability considerations (see Figure 2 for Domain headings).^29^ Domains were scored between 0 (not at all) and 3 (to a large extent) by consensus and summarised via radar plot.

**Figure 2.**
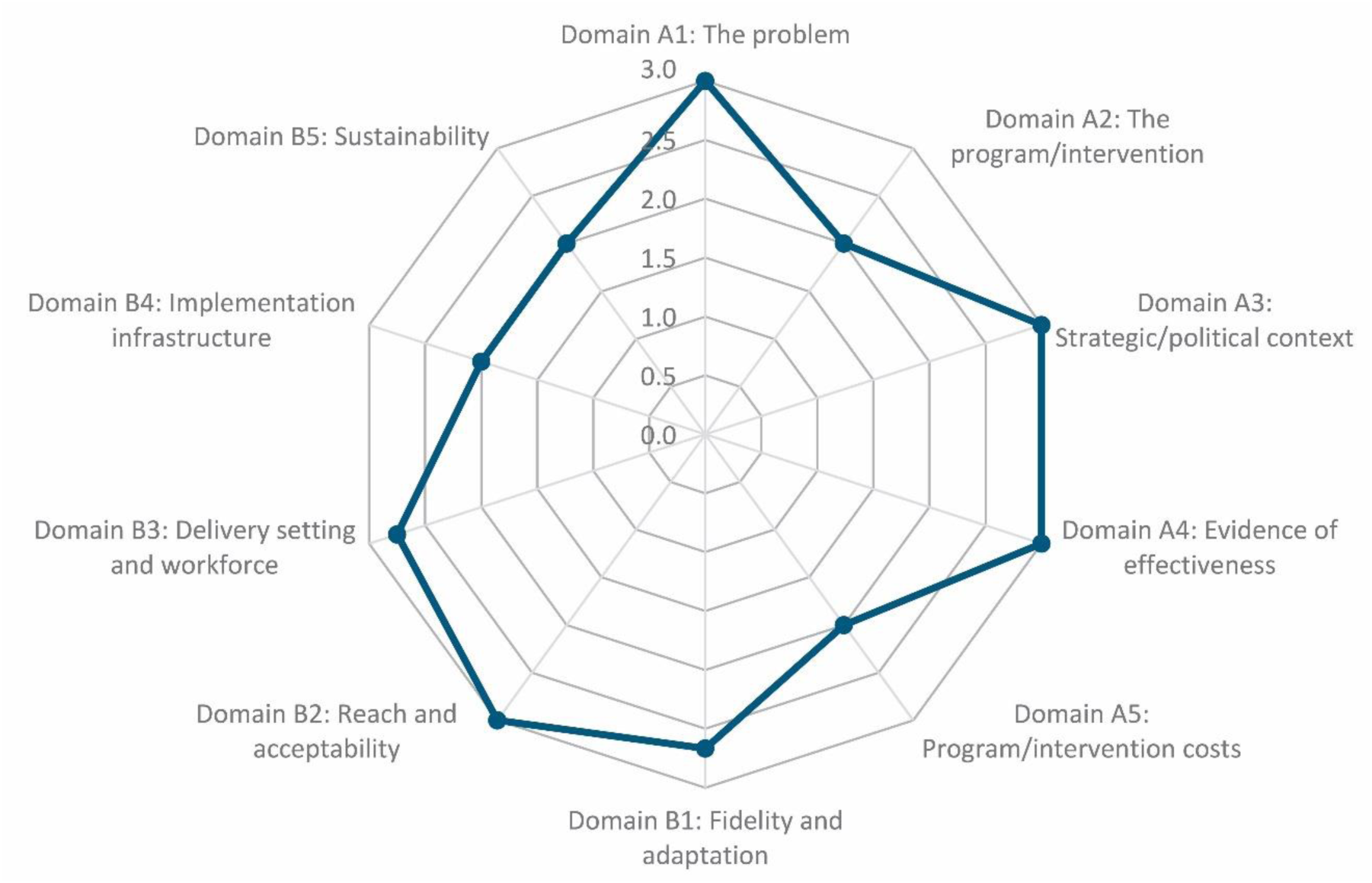
Intervention Scalability Assessment Tool (ISAT) radar plot depicting factors influencing Region 2’s readiness for NROL implementation. Domains A1-5 focus on background contextual information and B1-5 on implementation and sustainability. Each domain is scored for readiness between 0 (not at all) to 3 (to a large extent), therefore facilitators are depicted nearer the outside of the circle and barriers more centrally.

In Step 4, the TIDieR-Rehab checklist,^18^ with the Telehealth extension,^19^ was used to report the adapted innovation. The TIDieR-Rehab extends the original TIDieR checklist^17^ to better capture the complexity of rehabilitation interventions. Content for the TIDieR-Rehab checklist was systematically updated from Region 1’s TIDieR checklist,^6^ with region specific content (new/adapted/removed) clearly marked.

## RESULTS

### Step 1: Assess rationale for intervention, and consider intervention-context fit of existing innovation

#### Establish rationale, and consider identified innovation and its core components

Region 2 faced persistent challenges in delivering the recommended levels of stroke rehabilitation, compounded by workforce shortages and fragmented pathways, and anticipated increased prevalence.^30^ Regional Stroke Rehab Leads from the region’s stroke delivery network identified NROL as a promising solution, informed by exposure at meetings, conferences and literature.

#### Map similarities/difference with original region and consider intervention-context fit

Table 1 shows the comparison of the similarities and differences between Region 1 (original) and Region 2 (new) and identifies the barriers and facilitators to NROL implementation. This work was led by coalition members: two designated Regional Stroke Rehab Leads (RW,LJ) from Region 2, and the NROL Operational Lead (AP) and Clinical-Academic staff (SA,LC) from Region 1.

**Table 1:**
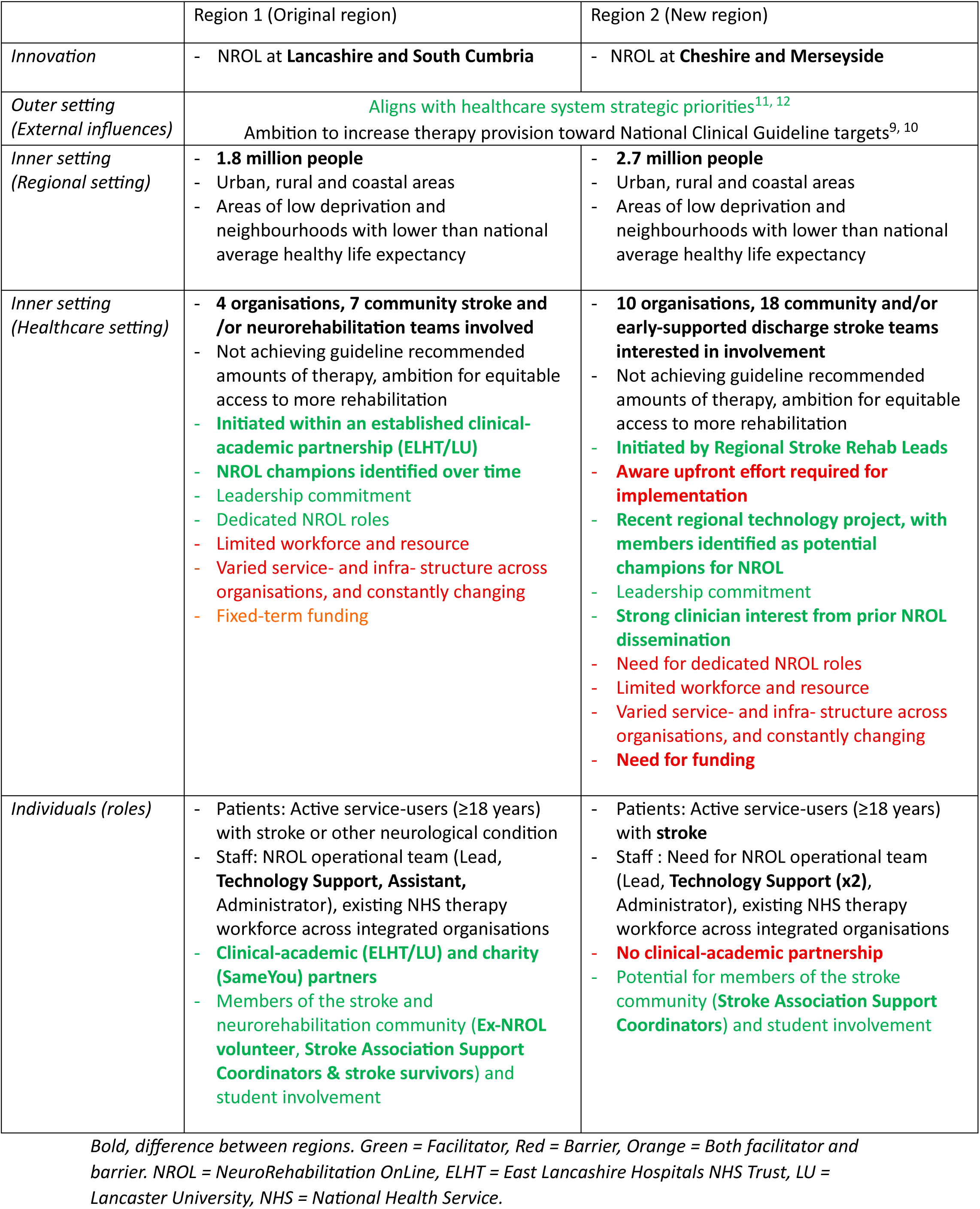
Comparison of contextual factors between original and new regions, including barriers and facilitators to NROL implementation

Key differences included that Region 2 was larger (population and number of organisations) and had a stroke-specific focus, with future ambition to extend to other neurological conditions. While most eligible organisations (10/14) showed interest in NROL, four opted not to participate due to competing priorities, or hesitancy around new initiatives and technology. Another difference was the absence of an established clinical-academic partnership for the project, contrasting Region 1 where such a partnership was central. Region 2, however, felt they benefitted from Regional Stroke Rehab Lead initiation and leadership, as they had existing knowledge of the ‘people and processes’ covering all organisations and recent technology project engagement, which could help expedite upfront implementation efforts (e.g. stakeholder engagement, governance approvals).

Key similarities included recognition that NROL had strong strategic alignment,^11, 12^ a significant facilitator. Both regions identified lack of workforce and resource, and varied service-and infra-structure across organisations as barriers for implementation. There was a preference to maintain dedicated NROL operational roles, however funding was needed to recruit staff. Two Technology Support roles were prioritised due to existing availability of therapy assistance within the workforce and the size of the region. Region 2 recognised NROL’s potential to optimise workforce use and drew on Region 1’s solutions to service challenges.

Figure 2 depicts the factors influencing Region 2’s context-specific readiness for NROL implementation. ISAT scoring showed NROL was seen as a valuable innovation, though not a panacea (A2). Funding needs (A5), infrastructure gaps (B4) and long-term sustainability were identified as areas for response, notably highlighting need for dedicated NROL operational staff.

### Step 2: Plan for and undertake adaptations

#### Accessing necessary resource

In response to the identified barriers, seed funding was secured through a national health funding body enabling recruitment of the dedicated NROL operational team (including Lead, AB) and a modest budget for equipment and materials.

#### Form adaptation team and create learning collaborative

An adaptation team was formed comprising Regional Stroke Rehab Leads (n = 2, Region 2), NROL Operational teams (n = 8, both regions), and Clinical-Academic staff (n = 2, Region 1). The adaptation team met regularly to iteratively adapt NROL for Region 2. Adaptations were informed by Step 1, ongoing regional, organisational and service needs, and stakeholder feedback. The new region viewed the involvement of the original region positively.

A regional NROL learning collaborative was established, and was iterated over time, to support shared decision-making and co-production. This included existing workforce (regional technology project members, therapy staff, local administrators and decision-makers), patients, and members of the stroke community (Stroke Association Support Coordinators) from Region 2, alongside the adaptation team and Region 1’s Stroke and Neurorehabilitation Service Manager (HV).

#### Adapt innovation, materials and procedures

Adaptations were planned and undertaken to achieve the best fit of the NROL core and peripheral components within Region 2, and key components are summarised in Figure 3. The remaining peripheral components were all maintained and are described in the TIDieR-Rehab checklist (Supplementary file). Responsive adaptations are detailed in Step 3.

**Figure 3:**
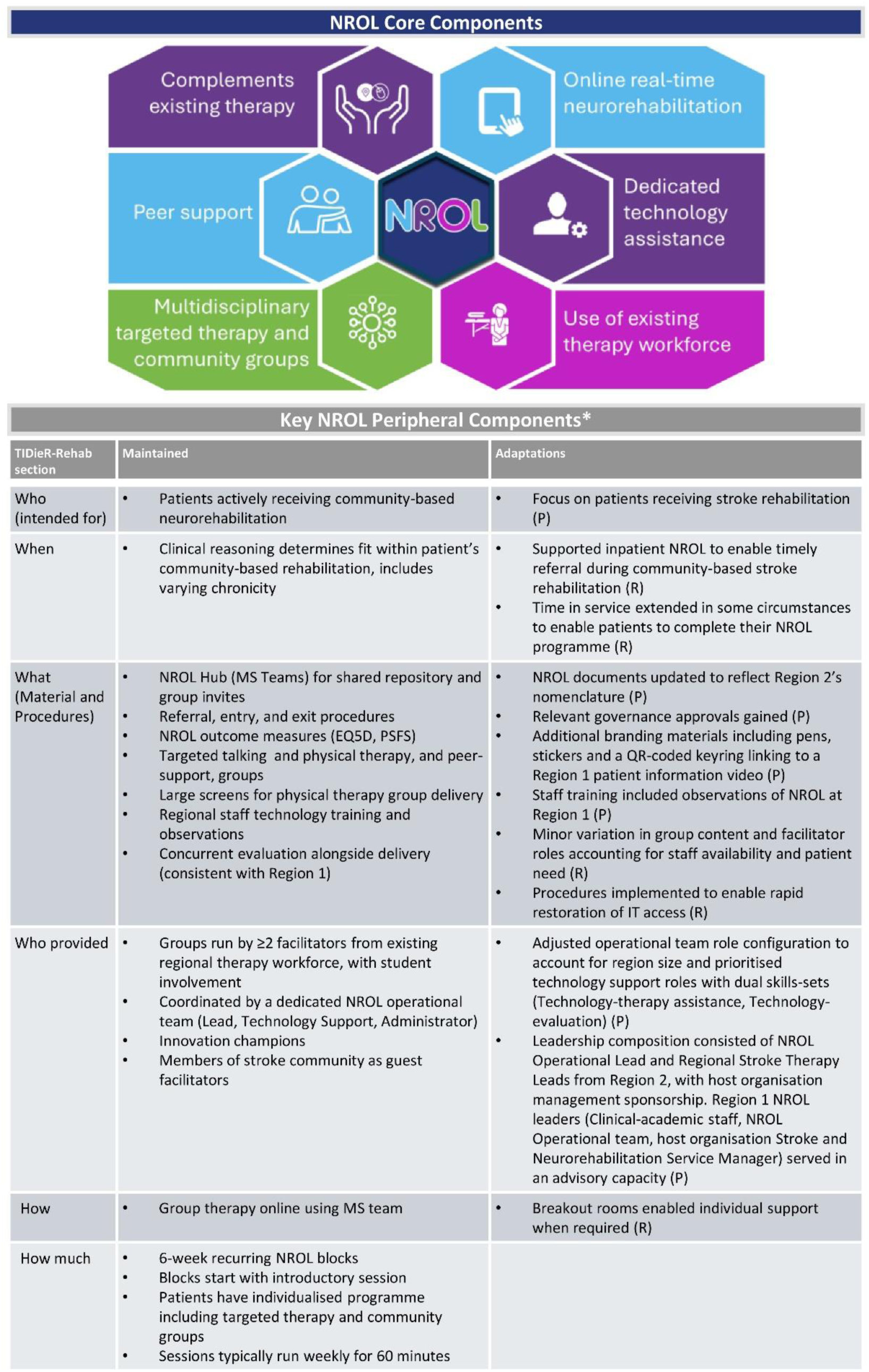
Core and key peripheral components for NROL adapted for Region 2. Key peripheral components are detailed under TIDieR Rehab sections. Planned adaptations (P) determined during ADAPT step 2 and responsive adaptations (R) during ADAPT step 3. *Remaining peripheral components were maintained and are detailed in the TIDieR-Rehab checklist (supplementary file).

All NROL core components were retained. The description of one core component was modified to clarify the model was intended for use in ‘neurorehabilitation’ rather than specifying stroke and neuro-rehabilitation.

Key NROL peripheral components were aligned with the relevant TIDieR-Rehab section headings, and categorised as either ‘maintained’ or ‘adaptations’. The basic structure of NROL was maintained (i.e. How/How much). Planned adaptations were made to the ‘Who’ NROL was intended for and ‘Who provided’, and included offering NROL only to patients receiving stroke rehabilitation, and adjustment to operational team and leadership role configuration. Planned adaptations were co-produced to ‘What’ materials with the learning collaborative, including updating key documents (shared by Region 1), gaining relevant governance approvals, and expanding NROL visibility with additional branding materials.

#### Recruit staff to deliver innovation, identify and prepare champions

Region 2 NROL leaders in collaboration with the learning collaborative, recruited NROL champions and group facilitators from high-readiness organisations. Innovation champions were key in supporting governance, advocacy, and troubleshooting. Facilitators received targeted training, including technology and procedural sessions, and Region 1 observations. Meetings were held for delivery staff, champions, and leaders, with updates shared to wider teams.

### Step 3: Plan for and undertake piloting and evaluation of the adapted innovation

#### Ensure readiness

Innovation readiness was confirmed by the Region 2 NROL operational team and champions once the infrastructure, training, equipment, regional approvals and staffing were in place. Organisational readiness required local approvals, staff training and baseline service data to provide a reference point for evaluation.

#### Conduct small tests of change

Pilot testing was initially conducted at group-level through two pilot groups: a fatigue management group and a speech and language therapy group. These allowed assessment of acceptability and practicality. Staff and patient feedback informed iterative refinements to processes, logistics and group content.

#### Phased integration of interested organisations

Following successful group-level piloting, block-level piloting began. This involved phased integration of organisations with high-readiness into 6-week NROL blocks running seven groups (Cognitive education, Emotional adjustment, Dysarthria, Aphasia, Balance and Mobility, Upper limb, Community peer-support).

All interested organisations were integrated. Seven organisations (8 teams) adopted the programme initially, one further organisation (3 teams) was added in the second block and two more (2 teams) in the third. Five teams remained pending. The learning collaborative expanded to ensure regional representation.

Evaluation ran alongside implementation and centred on reviewing key peripheral components, using routinely collected clinical and service-delivery metrics and stakeholder feedback and was consistent with Region 1 evaluation. Region 2 NROL leaders co-produced summaries for stakeholders, including Region 1.

#### Document and classify responsive adaptations

Regular review meetings allowed iterative tailoring. Responsive adaptations are summarised in Figure 3 and included in the TIDieR-Rehab checklist (Supplementary file). Key responsive adaptations included tailoring to fit within the pathway such as supported inpatient participation and extended time in the community-based service when appropriate. Existing workforce influenced group content and facilitator roles (e.g. Allied health staff trained to lead well-being group). IT access issues prompted new resolution procedures, and breakout rooms (new functionality) were introduced, with procedures shared with Region 1.

### Step 4: Implement and maintain the adapted innovation

#### Conduct ongoing training

Findings from pilot testing and evaluation informed ongoing training. Staff were encouraged to observe live NROL sessions, in both regions, to support referral decisions and experiential learning. Cross-region observation fostered shared learning and networking. New staff were introduced to NROL during orientation. New group facilitators received targeted training in both technology and procedures. General information about NROL was regularly disseminated by local and regional NROL staff and champions.

#### Consider sustainment

Sustainment was considered from the out-set and involved continued dialogue with wide stakeholders, including decision-makers. Presentations were made to organisation transformation and improvement leaders. Advantages of the programme were identified and included staffing efficiencies, increased therapy contact, and savings in travel time, cost, and carbon emissions. A co-produced blueprint outlined full implementation and evaluation plans. Further fixed-term funding was secured, providing time to develop a business case for long-term regional investment. NROL delivery is ongoing.

#### Report adapted innovation described using TIDieR-Rehab checklist

The adapted NROL model for Region 2 was documented using the TIDieR-Rehab checklist, updated from Region 1’s version^6^ to reflect adaptations (see supplementary file). This co-production process involved the Region 2 learning collaborative to ensure accuracy and resonance.

## DISCUSSION

This study illustrates how the NROL telerehabilitation model was adapted for a new region using a structured, co-produced process. Core components were retained, while adaptations tailored role configuration, materials and fit within service pathways. The ADAPT guidance alongside implementation science knowledge provided a systematic approach to planning, piloting, implementing, and reporting the adapted model.

The adaptation process was initiated and led by Region 2 Stroke Rehab leaders who had relationships with local organisations and deep community knowledge facilitating implementation efforts. Involving the original region (Region 1) accelerated implementation by transferring tacit knowledge, tested materials, and credibility. While the ADAPT guidance warns working with innovation developers could risk fixed views or power imbalances,^16^ this did not materialise. The new region viewed the involvement positively, likely due to co-production principles, open dialogue, and mutual recognition. This approach provided a trusted environment to explore viable adaptations to meet local needs.

Creative solutions were fostered. Adjustments to role configurations and recruitment reflected the wider workforce landscape and leveraged diverse skill-sets. For example, Region 2 prioritised recruitment for two NROL technology support roles. The opportunity was harnessed to establish a patient-facing technology job creation, and capitalised on dual skill-sets (e.g. assistant workforce), that could help sustain digital practice. Another example, in response to a system-wide lack of psychology staff, involved allied health staff being trained to deliver a well-being group with oversight from a clinical psychologist. This role agility aligns with the recent UK stroke guideline emphasis on what needs to be done rather than who does it.^9^ Creative solutions were also found for promoting NROL access, such as optimising fit within the service pathways (e.g. early exposure through inpatient participation) and introducing new materials (e.g. QR-coded keyring to share a patient information video).^31^ It may be that these adaptations actively enhanced joint decision-making between therapist and patient about service delivery preferences.^32^

Frameworks were applied selectively to help navigate the complexities of transferring across regions. The ADAPT guidance structured the adaptation process; CFIR and ISAT supported contextual analysis; and TIDieR-Rehab enabled transparent reporting. The process was non-linear, with evolving priorities. While these tools added rigour, they also introduced complexity. Overlapping constructs and terminology created challenges, requiring interpretive effort.^23, 33^ Co-production, including clinical-academic involvement, helped translate conceptual guidance into practice-relevant actions.^24, 34, 35^ Insights are summarised in the “Key Considerations for Adaptation” box, which may be helpful for researchers and implementers seeking to scale telehealth models.

Key Considerations for Adaptation

- Embed co-production and shared learning: Engage stakeholders throughout.
- Retain core functions, use creative solutions to meet needs: Tailor staffing models, materials and fit within pathway.
- Use structured guidance flexibly: Apply frameworks to support planning and contextual fit.
- Anticipate complexity: Expect evolving roles and shifting priorities.
- Plan for sustainment early: Co-produce a blueprint and highlight efficiencies.

A limitation was that the project focused on a single adaptation case within well-connected national health system regions. Findings may not generalise to less resourced or less connected contexts. The use of embedded teams and qualitative feedback limits external validation but offers rich, practice-based insights. Further evaluation is needed to assess sustainability, cost-effectiveness, and equity.

This paper highlights the complexity and effort required to adapt and sustainably scale interventions, a challenge often under-recognised in healthcare and funding agendas. It showed the value of working across silos and with the original region to share information, evaluation and know-how, including with decision-makers. Working as a cross-regional learning health system provided a means to accelerate the integration of NROL in the new context.^36^ The resource demands of thorough adaptation underscore the need for sustainable, bottom-up funding models. The results affirm that strategic adaptation must address clinician, patient, service, and system-level factors, with clinical-academic partnerships a potential mechanism for translating frameworks into scalable, context-sensitive tools.^37^

## Conclusion

This study offers a transferable approach for scale-up, providing a real-world example of adapting a complex telerehabilitation model using structured implementation science knowledge. It offers practical insights into how core components can be maintained while adapting elements to new contexts. As digital models expand, structured and context-sensitive adaptation grounded in co-production and implementation science approaches, will be essential for sustainable scale-up across diverse health systems. These findings may inform broader efforts in digital transformation, workforce redesign, and service innovation beyond telerehabilitation.

## Data Availability

All data produced in the present study are available upon reasonable request to the authors

## ACKNOWLEDGMENTS

We gratefully acknowledge the contributions of staff, patients and carers from the participating stroke and neurorehabilitation services within the Cheshire and Merseyside and Lancashire and South Cumbria Integrated Care Systems, along with their affiliated Integrated Stroke and Neuro Delivery Networks and the Stroke Association. Our thanks also go to the charity SameYou for their ongoing partnership.

## STATEMENTS AND DECLARATIONS

### Ethical considerations

Not applicable.

### Consent to participate

Not applicable.

---

**Supplementary file 1: TIDieR-Rehab Checklist for NeuroRehabilitation OnLine (NROL) at Cheshire and Merseyside**

The TIDieR-Rehab checklist^1^ was used, informed by the Telehealth extension^2^, to report the adapted NROL innovation at Cheshire and Merseyside (new region, Region 2).

Content was systematically updated from the TIDieR checklist described for Lancashire and South Cumbria (original region, Region 1)^3^:

1. The TIDieR checklist content for NROL at Lancashire and South Cumbria was reallocated into the sections of the TIDieR-Rehab checklist.
2. Each TIDieR-Rehab checklist section was updated to reflect components maintained and adapted with edits clearly marked to enable comparison:
  - **Bold font** for new/adapted content for NROL at Cheshire and Merseyside
  - Strikethrough for removed content
  - *Italics for additions/amendments* relevant to both regions

**Supplementary Table 1:**
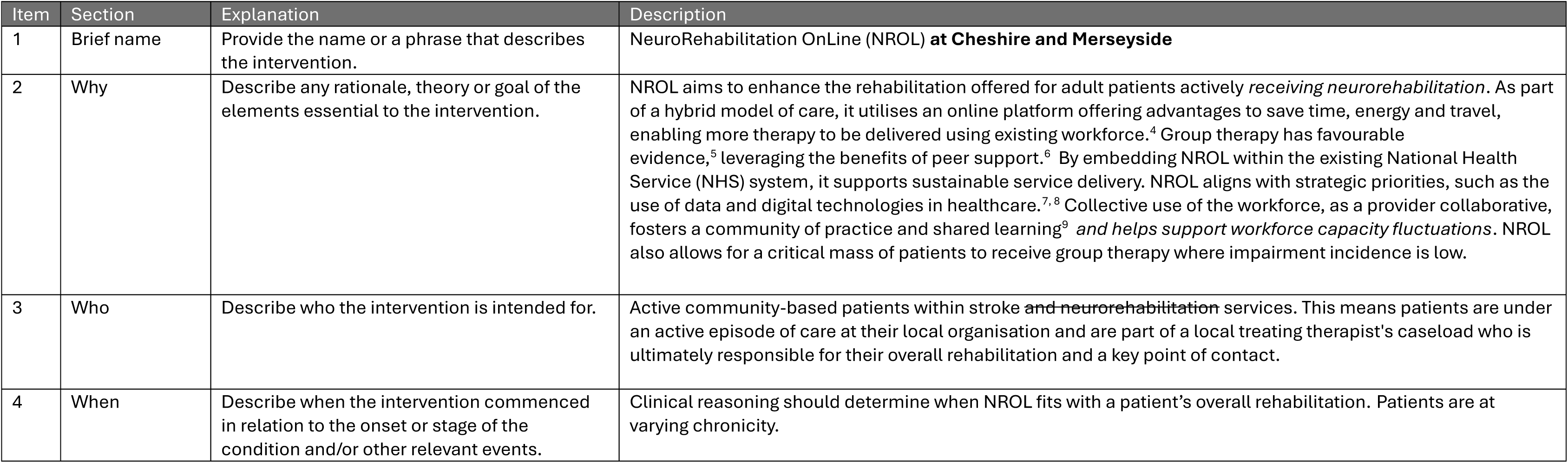
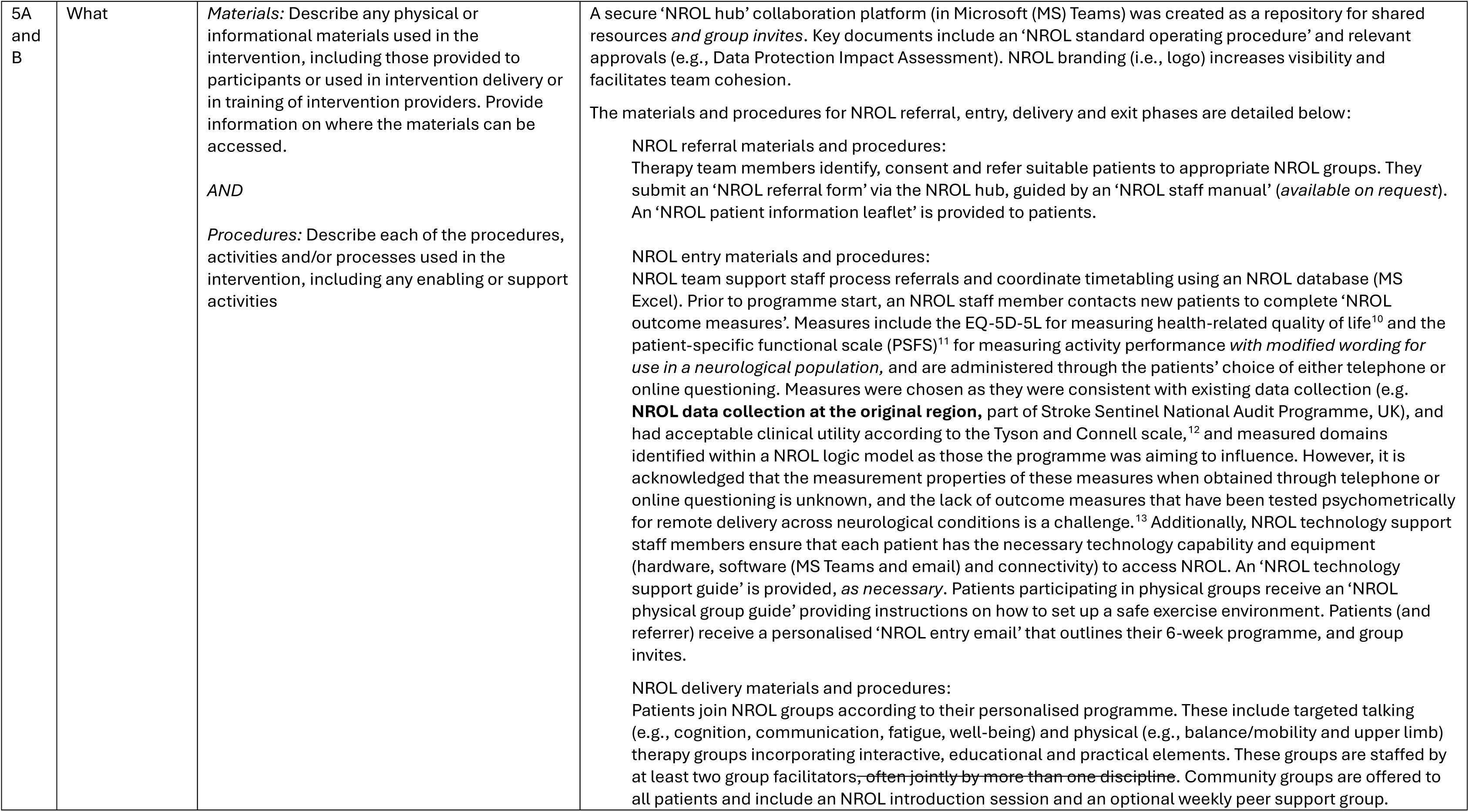
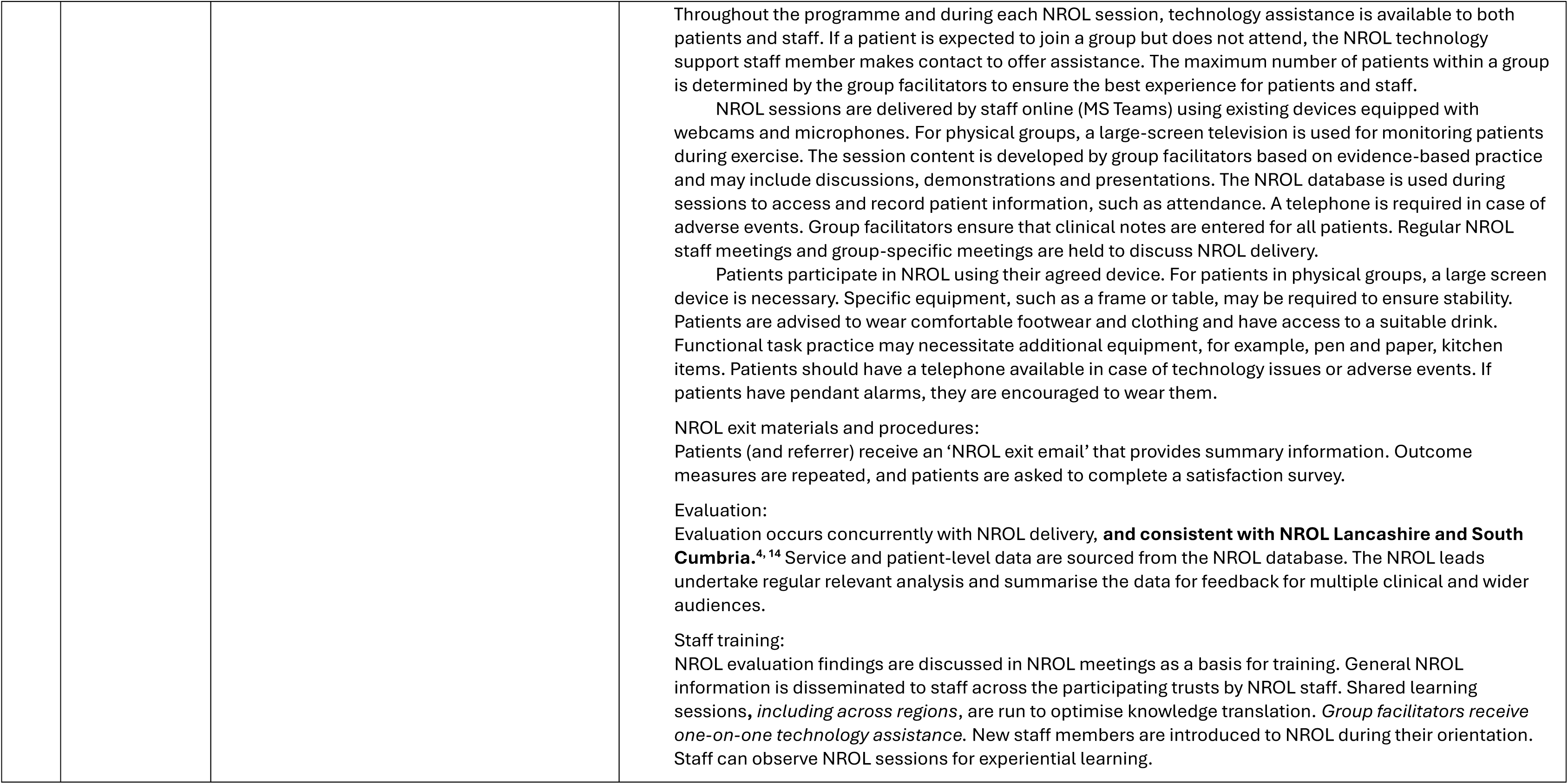
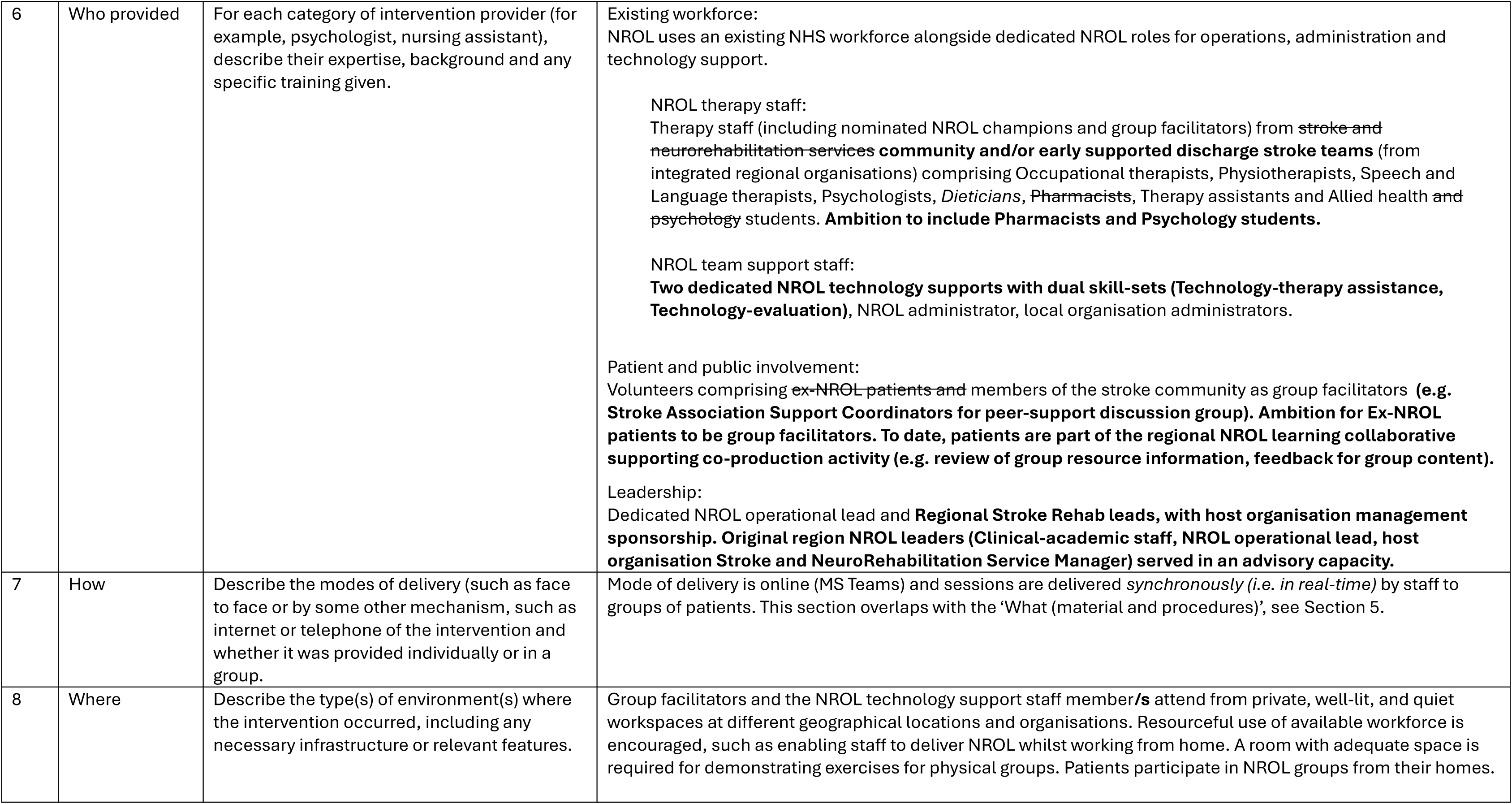
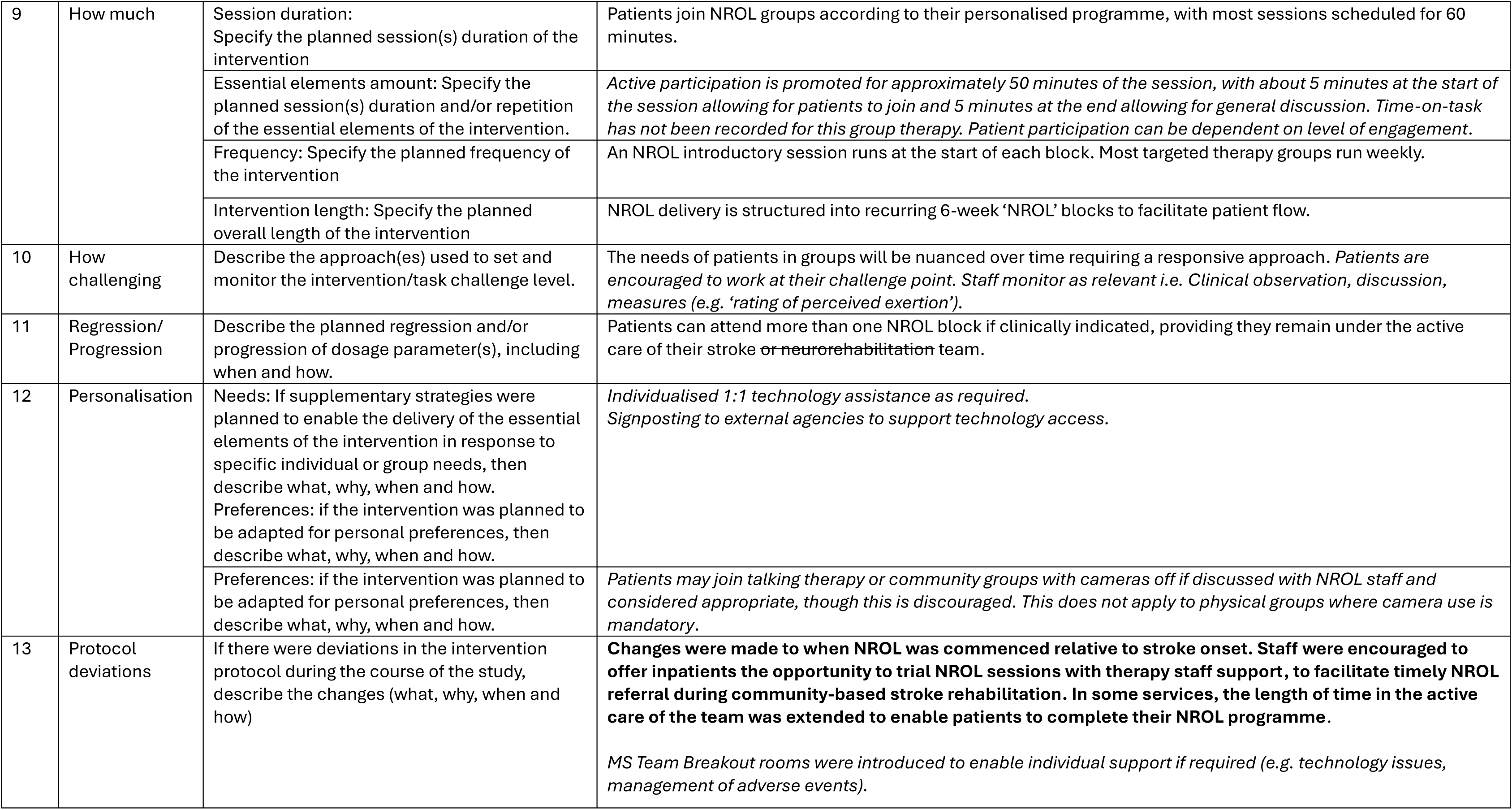
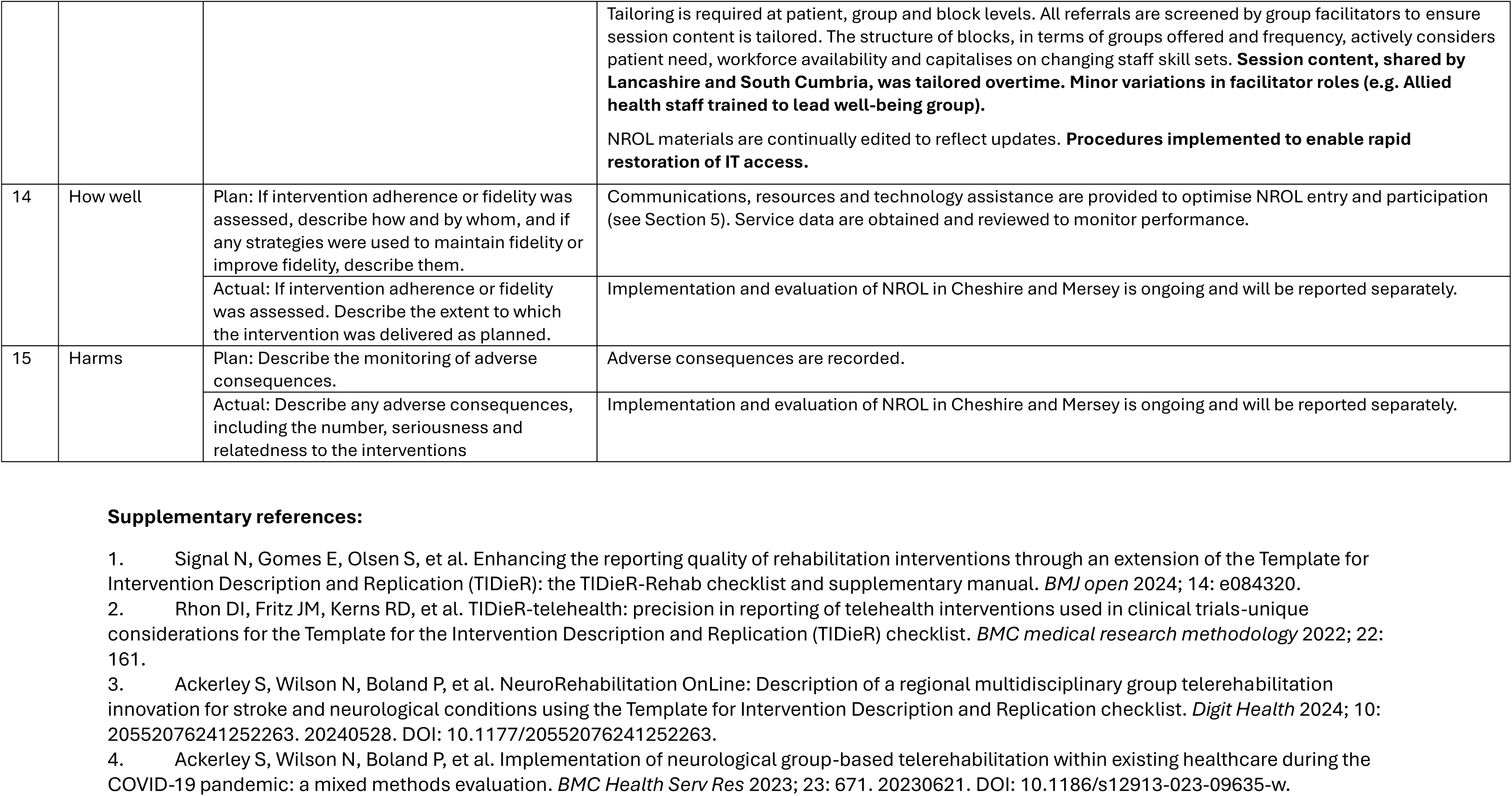
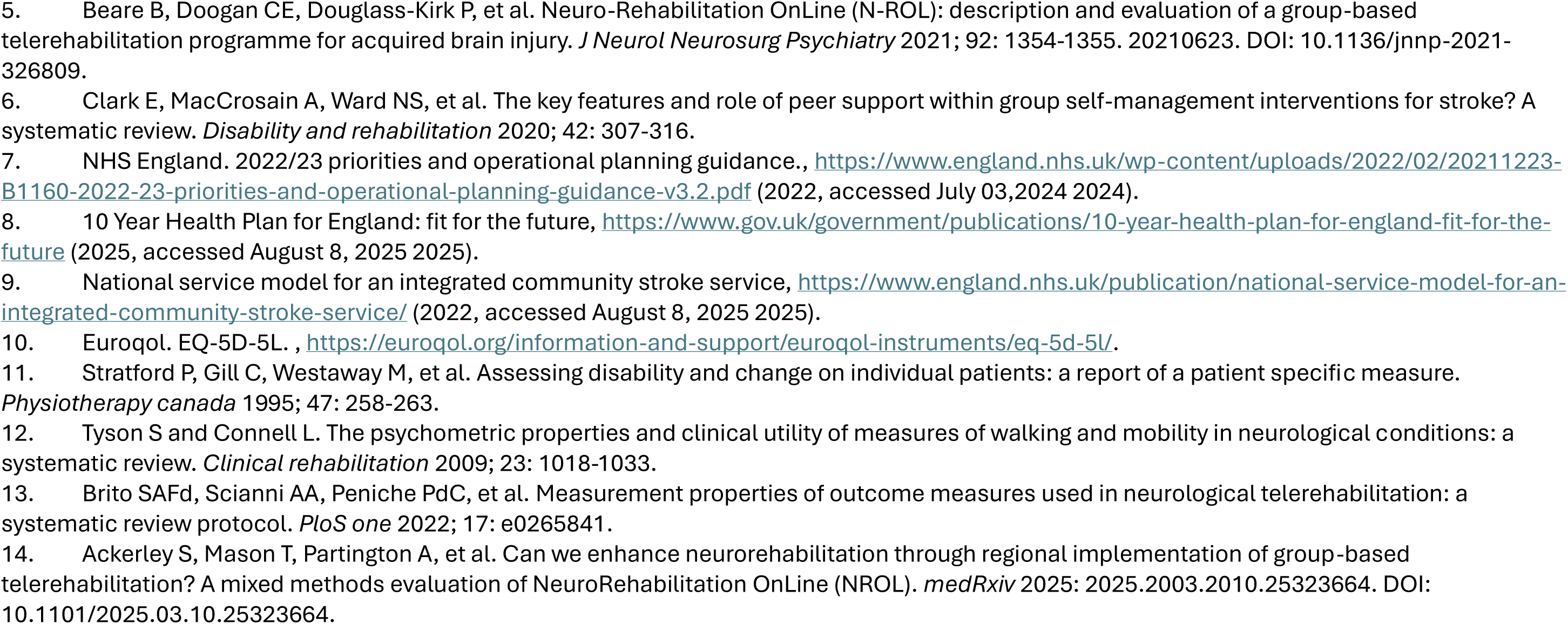
TIDieR-Rehab Checklist for NeuroRehabilitation OnLine (NROL) at Cheshire and Merseyside

## REFERENCES

1. Kings College London. Stroke Sentinel National Audit Programme (SSNAP). SSNAP Annual Report 2023., https://www.strokeaudit.org/Documents/National/Clinical/Apr2022Mar2023/Apr2022Mar2023-AnnualReport.aspx (2023, accessed July 3 2025).

2. Stroke Foundation. National Stroke Audit - Rehabilitation Services Report 2024. Melbourne, Australia., https://informme.org.au/media/e00n04xd/national-stroke-rehabilitation-services-report-2024-final-20-11-2024.pdf (2024, accessed July 4 2025).

3. Ontario OAGo. Value for Money Audit Cardiac Disease and Stroke Treatment., https://www.auditor.on.ca/en/content/annualreports/arreports/en21/AR_Cardiac_en21.pdf (2021, accessed July 4 2025).

4. Raymond MJ, Christie LJ, Kramer S, et al. Delivery of Allied Health Interventions Using Telehealth Modalities: A Rapid Systematic Review of Randomized Controlled Trials. In: Healthcare 2024, p.1217. Multidisciplinary Digital Publishing Institute.

5. Laver KE, Adey-Wakeling Z, Crotty M, et al. Telerehabilitation services for stroke. Cochrane Database Syst Rev 2020; 1: CD010255. 20200131. DOI: 10.1002/14651858.CD010255.pub3.

6. Ackerley S, Wilson N, Boland P, et al. NeuroRehabilitation OnLine: Description of a regional multidisciplinary group telerehabilitation innovation for stroke and neurological conditions using the Template for Intervention Description and Replication checklist. Digit Health 2024; 10: 20552076241252263. 20240528. DOI: 10.1177/20552076241252263.

7. Ackerley S, Wilson N, Boland P, et al. Implementation of neurological group-based telerehabilitation within existing healthcare during the COVID-19 pandemic: a mixed methods evaluation. BMC Health Serv Res 2023; 23: 671. 20230621. DOI: 10.1186/s12913-023-09635-w.

8. Ackerley S, Mason T, Partington A, et al. Can we enhance neurorehabilitation through regional implementation of group-based telerehabilitation? A mixed methods evaluation of NeuroRehabilitation OnLine (NROL). medRxiv 2025: 2025.2003.2010.25323664. DOI: 10.1101/2025.03.10.25323664.

9. Royal College of Physicians. National Clinical Guideline for Stroke for the United Kingdom and Ireland. , https://www.strokeguideline.org/app/uploads/2023/04/National-Clinical-Guideline-for-Stroke-2023.pdf (2023, accessed July 1 2024).

10. National Institutes for Health and Care Excellence (NICE). Stroke rehabilitation in adults. NICE guideline [NG236], https://www.nice.org.uk/guidance/ng236 (2023, accessed July 4 2025).

11. National service model for an integrated community stroke service, https://www.england.nhs.uk/publication/national-service-model-for-an-integrated-community-stroke-service/ (2022, accessed August 8 2025).

12. NHS England. The NHS Long Term Plan., https://webarchive.nationalarchives.gov.uk/ukgwa/20230418155402/ https://www.longtermplan.nhs.uk/publication/nhs-long-term-plan/ (2019, accessed September 15 2025).

13. 10 Year Health Plan for England: fit for the future, https://assets.publishing.service.gov.uk/media/6888a0b1a11f859994409147/fit-for-the-future-10-year-health-plan-for-england.pdf (2025, accessed August 8 2025).

14. World Health Organization. Nine steps for developing a scaling-up strategy. Geneva: WHO ExpandNet., https://iris.who.int/bitstream/handle/10665/44432/9789241500319_eng.pdf?sequence=1 (2010, accessed August 15 2025).

15. Uddin J, Joshi VL, Wells V, et al. Adaptation of complex interventions for people with long-term conditions: a scoping review. Translational Behavioral Medicine 2024; 14: 514–526.

16. Moore G, Campbell M, Copeland L, et al. Adapting interventions to new contexts-the ADAPT guidance. BMJ 2021; 374: n1679. 20210803. DOI: 10.1136/bmj.n1679.

17. Hoffmann TC, Glasziou PP, Boutron I, et al. Better reporting of interventions: template for intervention description and replication (TIDieR) checklist and guide. Bmj 2014; 348.

18. Signal N, Gomes E, Olsen S, et al. Enhancing the reporting quality of rehabilitation interventions through an extension of the Template for Intervention Description and Replication (TIDieR): the TIDieR-Rehab checklist and supplementary manual. BMJ open 2024; 14: e084320.

19. Rhon DI, Fritz JM, Kerns RD, et al. TIDieR-telehealth: precision in reporting of telehealth interventions used in clinical trials-unique considerations for the Template for the Intervention Description and Replication (TIDieR) checklist. BMC medical research methodology 2022; 22: 161.

20. Van Campe F, Chambaere K, Pivodic L, et al. Systematic adaptation of public health palliative care interventions across settings using ADAPT guidance: Methodological learnings from the EU NAVIGATE project. Palliative Medicine 2025; 39: 460–472.

21. Connell L, Ackerley S and Rycroft-Malone J. Applying the updated MRC framework for developing and evaluating complex interventions with integrated implementation conceptual knowledge: an example using NeuroRehabilitation OnLine. Frontiers in Health Services 2025; 5: 1562627.

22. Damschroder LJ, Reardon CM, Widerquist MAO, et al. The updated Consolidated Framework for Implementation Research based on user feedback. Implement Sci 2022; 17: 75. 20221029. DOI: 10.1186/s13012-022-01245-0.

23. Skivington K, Matthews L, Simpson SA, et al. A new framework for developing and evaluating complex interventions: update of Medical Research Council guidance. BMJ 2021; 374: n2061. 20210930. DOI: 10.1136/bmj.n2061.

24. Rycroft-Malone J, Graham ID, Kothari A, et al. Research Coproduction: An Underused Pathway to Impact. International Journal of Health Policy and Management 2024.

25. Hickey G, Brearley S, Coldham T, et al. Guidance on co-producing a research project. 2018. Southampton: INVOLVE 2021.

26. Hoekstra F, Mrklas K, Khan M, et al. A review of reviews on principles, strategies, outcomes and impacts of research partnerships approaches: a first step in synthesising the research partnership literature. Health Research Policy and Systems 2020; 18: 1–23.

27. Powell BJ, Waltz TJ, Chinman MJ, et al. A refined compilation of implementation strategies: results from the Expert Recommendations for Implementing Change (ERIC) project. Implement Sci 2015; 10: 21. 20150212. DOI: 10.1186/s13012-015-0209-1.

28. Waltz TJ, Powell BJ, Fernandez ME, et al. Choosing implementation strategies to address contextual barriers: diversity in recommendations and future directions. Implement Sci 2019; 14: 42. 20190429. DOI: 10.1186/s13012-019-0892-4.

29. Milat A, Lee K, Conte K, et al. Intervention Scalability Assessment Tool: a decision support tool for health policy makers and implementers. Health research policy and systems 2020; 18: 1.

30. King D, Wittenberg R, Patel A, et al. The future incidence, prevalence and costs of stroke in the UK. Age and ageing 2020; 49: 277–282.

31. NROL Patient Introduction, https://www.youtube.com/watch?v=rdek3xrkKBg (2024, accessed September 17 2025).

32. Cook R, Haydon HM, Thomas EE, et al. Digital divide or digital exclusion? Do allied health professionals’ assumptions drive use of telehealth? Journal of Telemedicine and Telecare 2025; 31: 376–385.

33. Nilsen P. Making sense of implementation theories, models, and frameworks. Implementation Science 30. Springer, 2020, pp.53–79.

34. Kislov R, Wilson P and Boaden R. The ‘dark side’of knowledge brokering. Journal of health services research & policy 2017; 22: 107–112.

35. Metz A, Jensen T, Farley A, et al. Is implementation research out of step with implementation practice? Pathways to effective implementation support over the last decade. Implementation research and practice 2022; 3: 26334895221105585.

36. Foy R, Carder P, Johnson S, et al. What should a learning health system look like? BMJ Open Quality 2025; 14.

37. Thomas EE, Taylor ML, Ward EC, et al. Beyond forced telehealth adoption: a framework to sustain telehealth among allied health services. Journal of Telemedicine and Telecare 2024; 30: 559–569

## Supplementary References

1. Signal N, Gomes E, Olsen S, et al. Enhancing the reporting quality of rehabilitation interventions through an extension of the Template for Intervention Description and Replication (TIDieR): the TIDieR-Rehab checklist and supplementary manual. BMJ open 2024; 14: e084320.

2. Rhon DI, Fritz JM, Kerns RD, et al. TIDieR-telehealth: precision in reporting of telehealth interventions used in clinical trials-unique considerations for the Template for the Intervention Description and Replication (TIDieR) checklist. BMC medical research methodology 2022; 22: 161.

3. Ackerley S, Wilson N, Boland P, et al. NeuroRehabilitation OnLine: Description of a regional multidisciplinary group telerehabilitation innovation for stroke and neurological conditions using the Template for Intervention Description and Replication checklist. Digit Health 2024; 10: 20552076241252263. 20240528. DOI: 10.1177/20552076241252263.

4. Ackerley S, Wilson N, Boland P, et al. Implementation of neurological group-based telerehabilitation within existing healthcare during the COVID-19 pandemic: a mixed methods evaluation. BMC Health Serv Res 2023; 23: 671. 20230621. DOI: 10.1186/s12913-023-09635-w.

5. Beare B, Doogan CE, Douglass-Kirk P, et al. Neuro-Rehabilitation OnLine (N-ROL): description and evaluation of a group-based telerehabilitation programme for acquired brain injury. J Neurol Neurosurg Psychiatry 2021; 92: 1354–1355. 20210623. DOI: 10.1136/jnnp-2021-326809.

6. Clark E, MacCrosain A, Ward NS, et al. The key features and role of peer support within group self-management interventions for stroke? A systematic review. Disability and rehabilitation 2020; 42: 307–316.

7. NHS England. 2022/23 priorities and operational planning guidance., https://www.england.nhs.uk/wp-content/uploads/2022/02/20211223-B1160-2022-23-priorities-and-operational-planning-guidance-v3.2.pdf (2022, accessed July 03,2024 2024).

8. 10 Year Health Plan for England: fit for the future, https://www.gov.uk/government/publications/10-year-health-plan-for-england-fit-for-the-future (2025, accessed August 8, 2025 2025).

9. National service model for an integrated community stroke service, https://www.england.nhs.uk/publication/national-service-model-for-an-integrated-community-stroke-service/ (2022, accessed August 8, 2025 2025).

48. Euroqol. EQ-5D-5L. , https://euroqol.org/information-and-support/euroqol-instruments/eq-5d-5l/.

11. Stratford P, Gill C, Westaway M, et al. Assessing disability and change on individual patients: a report of a patient specific measure. Physiotherapy canada 1995; 47: 258–263.

12. Tyson S and Connell L. The psychometric properties and clinical utility of measures of walking and mobility in neurological conditions: a systematic review. Clinical rehabilitation 2009; 23: 1018–1033.

13. Brito SAFd, Scianni AA, Peniche PdC, et al. Measurement properties of outcome measures used in neurological telerehabilitation: a systematic review protocol. PloS one 2022; 17: e0265841.

14. Ackerley S, Mason T, Partington A, et al. Can we enhance neurorehabilitation through regional implementation of group-based telerehabilitation? A mixed methods evaluation of NeuroRehabilitation OnLine (NROL). medRxiv 2025: 2025.2003.2010.25323664. DOI: 10.1101/2025.03.10.25323664.

